# Benchmarking the CoVID-19 pandemic across countries and states in the U.S.A. under heterogeneous testing

**DOI:** 10.1101/2020.05.01.20087882

**Authors:** Kenzo Asahi, Eduardo A. Undurraga, Rodrigo Wagner

**Author notes:** Address for correspondence: Eduardo A. Undurraga; Av. Vicuña Mackenna 4860, Macul CP 7820436, Santiago, Región Metropolitana, Chile.

## Abstract

Public health officials need to make urgent decisions to reduce the potential impact of the CoVID-19 pandemic. Benchmarking based on the increase in total cases or case fatality rates is one way of comparing performance across countries or territories (such as states in the USA), and could inform policy decisions about COVID-19 mitigation strategies. But comparing cases and fatality across territories is challenging due to heterogeneity in testing and health systems. We show two complementary ways of benchmarking across countries or US states. First, we used multivariate regressions to estimate the test-elasticity-of-COVID-19-case-incidence. We found a 10% increase in testing yielded ~9% (95% CI:4.2–3.4%; p<0.001) increase in reported cases across countries, and ~2% (95%CI:0.1-3.4%; p=0.03) increase across US states during the week ending April 10th, 2020. We found comparable negative elasticities for fatality rates (across countries: β =-0.77, 95%CI:-1.40– -0.14; p=0.02; US states: β=-0.15, 95%CI:-0.30-0.01; p=0.06). Our results were robust to various model specifications. Second, we decomposed the growth in cases into test growth and positive test ratio (PTR) growth to intuitively visualize the components of case growth. We hope these results can help support evidence-based decisions by public health officials as more consistent data hopefully becomes available.

## Introduction

As of April 18, 2020, SARS-CoV-2 has spread globally, resulting in 2.3 million reported infections and ~160,000 deaths in 185 countries and territories [1]. Substantial evidence shows the disease burden of CoVID-19 (illness caused by SARS-CoV-2) is higher than ascertained [2]. Public health officials need to make urgent decisions about interventions to reduce the potential impact of CoVID-19 with limited available evidence [3–5]. They have to continuously assess and adapt their decisions based on available resources, disease surveillance, and emerging scientific evidence.

Benchmarking based on the increase in total cases or case fatality rates is one way to compare performance across states, regions, or countries, and could inform policy decisions about mitigation strategies. But CoVID-19 case counts depend on lab-confirmation, so the number of reported cases is a function of testing [6]. Benchmarking may thus be of limited use because countries or even states within the USA do not have the same testing policies or test availability for SARS-CoV-2 [5]. Comparing fatality rates, which may be comparatively easier than comparing CoVID-19 incidence, is also challenging. To illustrate, the case fatality rate in China, adjusted for under-ascertainment, demography, and censoring (deaths lag infection), was estimated at ~1.4%. Still, the crude case fatality rate adjusted only by censoring was ~3.7% [7].

Ideally, public health officials would carry out testing based on representative random samples of the population (e.g., national, state) to estimate infection rates and case fatality rates. But countries and states have restricted testing considering its limited availability. To be sure, testing has increased globally [6]. But testing is far from random. Diagnostic tests are often used to allocate scarce healthcare resources, so tests target patients with more severe symptoms as CoVID-19 incidence increases. A purist would argue one cannot compare countries or territories with different levels of testing without random sampling. Acknowledging these limitations, we took a pragmatic approach to suggest two relatively simple ways of comparing territories, such as countries or US states, with varying degrees of testing. First, we used multivariate regressions to estimate the test-elasticity-of-CoVID-19-incidence, that is, the proportional increase in testing of CoVID-19 and in reported cases (both in logarithms). Our results were robust to various model specifications. Second, we decomposed the growth in cases into test growth and positive test ratio (PTR) growth to intuitively visualize the components of case growth. We hope our results can help public health officials when they need to benchmark vis-á-vis other territories.

## Materials and Methods

### Test-elasticity

We estimated the test-elasticity-of-CoVID-19-incidence for countries with CoVID-19 transmission globally (n=42; limited to countries that report the number of lab tests) [8] and for US states (n=51) [9]. We used a multivariate regression with the week-on-week change in the number of weekly lab-confirmed CoVID-19 cases or case fatality rates as our dependent variable. As independent variables, we used testing per capita, health expenditures, number of days since the 100^th^ lab-confirmed CoVID-19 case occurred, the share of the population ≥70 years of age with chronic respiratory disease. Variable definitions are shown in the Online Resource (Table S1).

We used a multivariate regression to estimate the test-elasticity-of-case-incidence. The regression specification is:

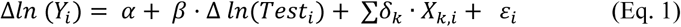

Where the subscript (*i*) stands for country or state, and (*k*) for covariates included. Because we used logarithms for *Y_i_* and *Test_i_, β* captures the test elasticity of CoVID-19 case incidence, controlling for a vector of covariates *X_k_* (Table S1). *ε* is an error term. Note that by first differencing both the dependent and the key variable, *β* accounts for any unobserved and time-invariant effect in a territory that may impact the level of testing and the level of the disease (e.g., income, quality of healthcare, age distribution). Still, we kept covariates as controls to test their impact on *β*.

### Case growth decomposition

A complementary way of benchmarking the evolution of cases vis-à-vis countries or US states with heterogeneous testing strategies is to use a simple algebraic decomposition. Because weekly cases can be decomposed as the multiplication of total tests *T_week_* and PTR *(Cases_week_/Tests_week_*), transforming to logarithms and taking the difference over weeks yields that the growth of cases can be decomposed as

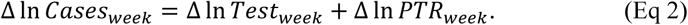

This growth decomposition is useful to visualize the source of change in reported cases for comparison across countries or US states.

## Results and discussion

### Test-elasticity

Figure 1 shows a “raw” (i.e., not controlling for covariates) estimate of the test-elasticity-of-cases, that is, the association between logarithmic changes in testing per capita and reported cases. Figure 1A shows a consistent association between the increase in testing per capita and reported CoVID-19 cases by country (β=0.86; 95%CI: 0.57–1.15; p<0.001). In other words, a 10% increase in testing was associated with a ~9 *%* increase in reported cases. Considering that a test elasticity=1 was included in the 95% confidence interval, we cannot reject that growth in weekly cases was fully proportional to increase in testing. By contrast, Figure 1B shows that for the US states, the test-elasticity of cases was substantially lower (β =0.17; 95%CI: 0.03–0.31; p=0.02), suggesting that the evolution of CoVID-19-case-growth in the US was largely driven by changes in PTR rather than testing. We get larger elasticities, more comparable to the global sample, when excluding outlier states in the USA and focusing on the same range of testing growth (β =0.54, p<0.0001, 95%CI: 0.36–0.71; Online Resource, Figure S1).

**Figure 1.**
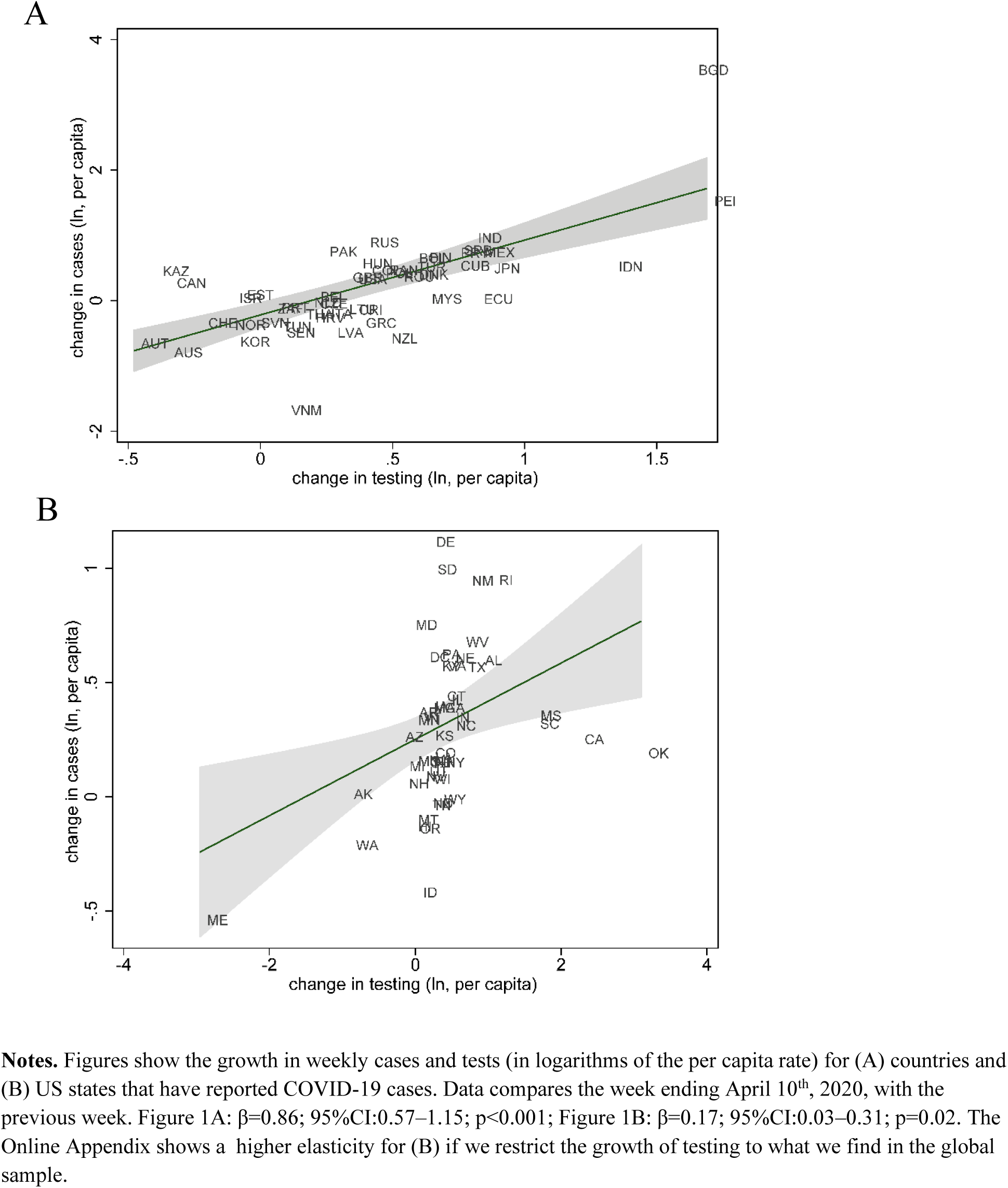
Changes in cases per capita relative to changes in the number of tests countries with reported COVID-19 cases (A), and for states in the USA (B).

Results in Table 1 are consistent with the findings in Figure 1A for the cross-country sample. With and without regression controls, we get point estimates for the elasticity around 0.85–0.88 (p-value <0.001; columns 1 through 3). A 10% increase in testing yielded about 9% (95% CI: 4.2–13.4%; p<0.001) increase in reported cases. Moreover, changes in testing explained about 40% of the variance in cases for our regressions (r^2^=0.41) and adding health expenditures, population, days since the 100^th^ CoVID-19 case, and share of the population ≥70 years of age with chronic respiratory disease added little explanatory strength to the model (r^2^=0.46). We obtained similar results for case fatality rate, but in the opposite direction (Table 1, columns 4 through 6). We found a test-elasticity-of-case-fatality slightly above minus one (β=-0.89; column 6). A 10% increase in testing yielded about 9% (95%CI: −12.2–5.6%; p<0.001) decrease in case fatality rates, with covariates adding limited explanatory strength (r^2^=0.45). To test robustness, we re-ran the regression with the most recent available data (April 11^th^ through April 17^th^); results were largely consistent. We found an elasticity for reported cases of β=0.77 (95%CI: 0.23-1.30; p=0.01; r^2^ =0.43, column 3), and β=-0.77 (95%CI:-1.40– −0.14; p=0.02, r^2^ =0.18, column 6) for case fatality rates (Online Resource, Table S2).

**Table 1.** Global regression estimates for the change in COVID-19 cases reported on the change in tests conducted by country in a week

|  | (1)<br>Cases<sup>a</sup> (ln) | (2)<br>Cases<sup>a</sup> (ln) | (3)<br>Cases<sup>a</sup> (ln) | (4)<br>Fatality<sup>b</sup> (ln) | (5)<br>Fatality<sup>b</sup> (ln) | (6)<br>Fatality<sup>b</sup> (ln) |
| --- | --- | --- | --- | --- | --- | --- |
| Testing <sup>c</sup> (ln) | 0.864***<br>(0.145) | 0.869***<br>(0.156) | 0.881***<br>(0.225) | -0.869***<br>(0.124) | -0.884***<br>(0.125) | -0.893***<br>(0.166) |
| Days since 100 cases <sup>d</sup> |  | 0.00115<br>(0.00719) | -0.0157<br>(0.0119) |  | -0.00419<br>(0.00716) | -0.0198<br>(0.0123) |
| Health expenditure per capita (USD, ln) |  |  | 0.148<br>(0.171) |  |  | 0.150<br>(0.132) |
| Aged 70+ with a chronic respiratory disease (ln) <sup>e</sup> |  |  | 0.0796<br>(0.134) |  |  | 0.0202<br>(0.195) |
| Population (ln) |  |  | 0.00573<br>(0.120) |  |  | 0.0696<br>(0.194) |
| Constant | -0.193*<br>(0.0912) | -0.225<br>(0.258) | -2.023<br>(1.526) | 0.607***<br>(0.0762) | 0.726**<br>(0.243) | -1.415<br>(1.852) |
| Observations | 42 | 42 | 42 | 38 | 38 | 38 |
| R-squared | 0.408 | 0.408 | 0.456 | 0.452 | 0.456 | 0.498 |

Table 2 shows results across states in the USA. As in Figure 1B, the magnitude of the elasticity was smaller for reported cases (β=0.18) and case fatality rate (β=-0.16). A 10% increase in testing yielded about 2% (95%CI: 0.1–3.4%; p=0.03, r^2^ =0.25, column 3) increase in reported cases, and ~2% (95%CI: −3.1–-0.2; p=0.03, r^2^ =0.14, column 6) decrease in case fatality rates (only significant with controls). R^2^ was smaller for regressions comparing US states than countries globally. Elasticities for US states using the most recent data available (April 11^th^ through April 17^th^) showed some qualitatively similar results. We found a test elasticity of β=0.17 (95%CI: 0.08-0.27; p<0.001, r^2^ =0.29, column 3) for reported cases, and β=-0.15 (95%CI: −0.30-0.01; p=0.06, r^2^ =0.12, column 6) for case fatality rates, though the latter was not significant at α=0.05 (Online Resource, Table S3).

**Table 2.** USA regression estimates for the change in COVID-19 cases reported by tests conducted by state in a week

|  | (1) | (2) | (3) | (4) | (5) | (6) |
| --- | --- | --- | --- | --- | --- | --- |
|  | Cases<sup>a</sup> (ln) | Cases<sup>a</sup> (ln) | Cases<sup>a</sup> (ln) | Fatality<sup>b</sup> (ln) | Fatality<sup>b</sup> (ln) | Fatality<sup>b</sup> (ln) |
| Testing <sup>c</sup> (ln) | 0.167*<br>(0.0700) | 0.169*<br>(0.0715) | 0.177*<br>(0.0810) | -0.0805<br>(0.0413) | -0.0832<br>(0.0467) | -0.164*<br>(0.0722) |
| Days since 100 cases <sup>d</sup> |  | -0.00897<br>(0.00750) | -0.0147<br>(0.00913) |  | 0.0147<br>(0.0120) | -0.0210<br>(0.0215) |
| Health expenditure per capita (USD, ln) |  |  | 0.643*<br>(0.274) |  |  | 0.00631<br>(0.403) |
| Aged 70+ with a chronic respiratory disease (ln) <sup>e</sup> |  |  | -0.0131<br>(0.182) |  |  | -0.298<br>(0.300) |
| Population (ln) |  |  | 0.0650<br>(0.0686) |  |  | 0.270<br>(0.166) |
| Constant | 0.252***<br>(0.0445) | 0.433*<br>(0.180) | -6.268<br>(3.177) | 0.434***<br>(0.0782) | 0.129<br>(0.296) | -4.311<br>(4.849) |
| Observations | 51 | 51 | 51 | 47 | 47 | 47 |
| R-squared | 0.154 | 0.177 | 0.250 | 0.0167 | 0.0430 | 0.140 |

**Notes**
\* $p < 0.05$ , \*\* $p < 0.01$ , \*\*\* $p < 0.001$ . Standard errors shown in parentheses. ln stands for natural logarithm.
<sup>a</sup> Confirmed cases of COVID-19<sup>2</sup> during the week ending April 10th, 2020.
<sup>b</sup> Case fatality rate is the ratio between deaths due to COVID-19 and cases in the same period.
<sup>c</sup> Testing is the amount of negative and positive tests informed to each country's health authority in the same period of cases [6].
<sup>d</sup> The number of days since the cumulated number of cases was equal or greater than 100 [6].
<sup>e</sup> Share over age 70 with a chronic respiratory disease is the share of the population with age equal or greater than 70 years old with a chronic respiratory disease, including chronic obstructive pulmonary disease and asthma.

### Case growth decomposition

Figure 2 shows a growth decomposition of CoVID-19 cases for countries (A) and US states (B). The origin (0, 0) shows both testing and PTR remained the same as the previous period. Figure 2 shows four quadrants where countries would fall when they increased or decreased in either testing or PTR. When both testing and PTR grow (quadrant I) or decrease (quadrant III), or countries o states move along the y-axis (change in PTR) or x-axis (change in tests), weekly change has a straightforward interpretation. But for quadrants II and IV, the net effect in cases is not obvious. To aid interpretation, we plotted a downward sloping line that represents the zero-case-growth (i.e., where Δln *Test_week_* + Δln *PTR_week_ =* 0; hence Δln *Test_week_ =* − Δln *PTR_week_*). Countries or states above the line increased case growth in the past week; those below the line have decreased case growth. Case growth in most countries moves in complex trajectories. The distance to the zero-case growth line is a visual clue for the overall increase in cases.

**Figure 2.**
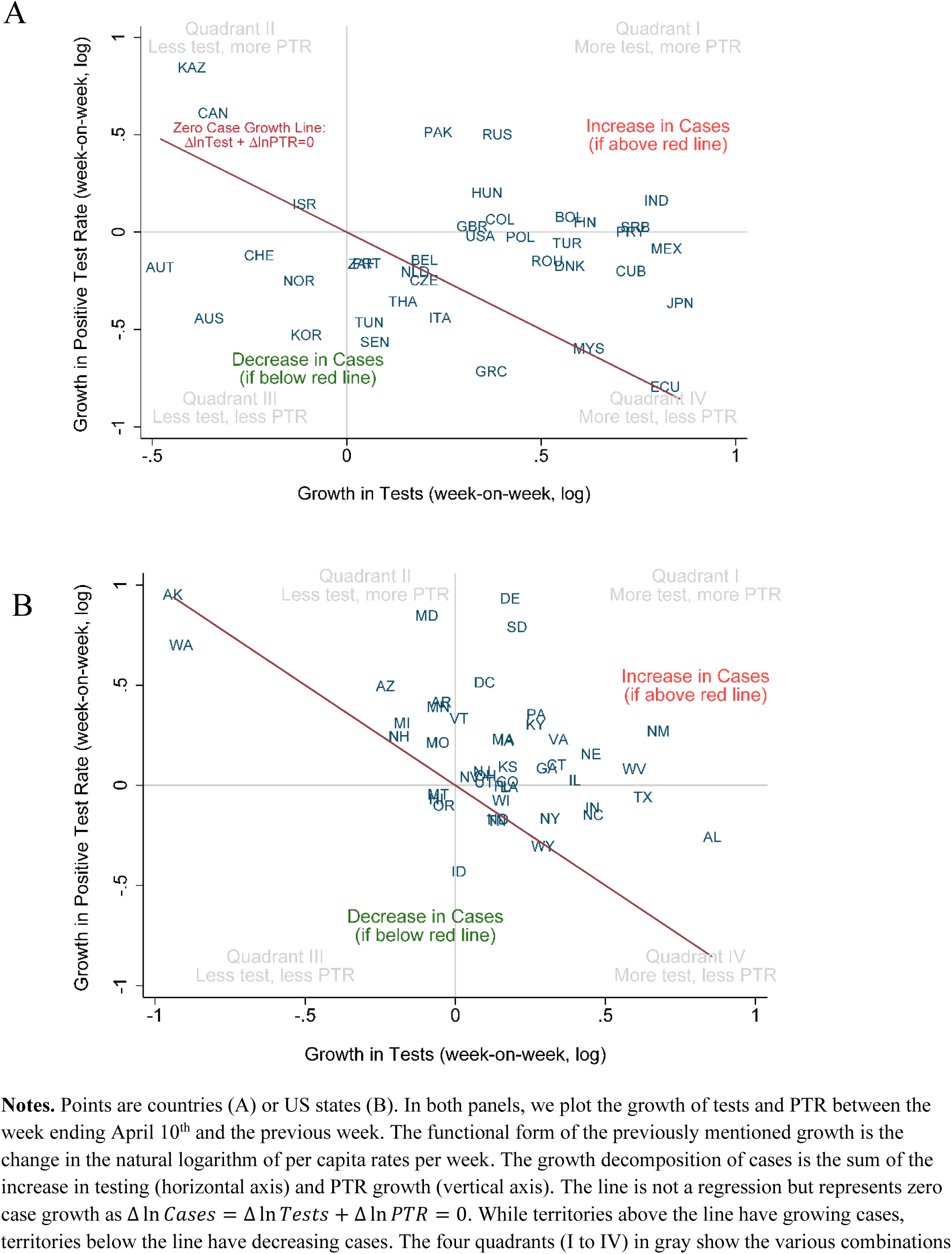
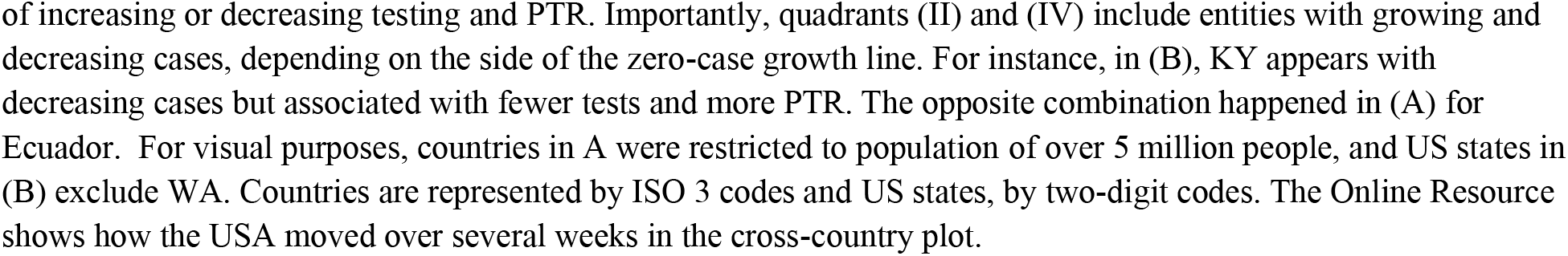
Mapping the Growth Decomposition of Cases: Testing Growth vs. PTR Growth. Week-on-week growth of Testing and PTR across (A) countries, and (B) US states.

In Figure 2A, we see that countries like Italy (ITA) and the United Kingdom (GBR) both increased the number of tests performed between April 10^th^ and April 17^th^. However, because PTR increased in the UK and decreased in Italy, we can conclude, solely by looking at Figure 2A, that CoVID-19 cases increased in the UK and decreased in Italy between April 3^rd^ and April 10^th^. On the other hand, countries like Ecuador (ECU) and Israel (ISR) appear on top of the line, meaning that growth in COVID-19 cases was approximately the same as in the previous week (Δ ln *Cases_week_ =* 0). Notably, this null change in case growth is accounted for by different levels of testing. In the graph, Israel decreased reported testing but increased the positive rate in the same proportion. In contrast, Ecuador massively increased testing during that week with a declining PTR, leaving case growth unchanged. This decomposition helps track case growth visually across jurisdictions with heterogeneous testing. Differences in testing need to be taken into account when benchmarking and communicating the evolution of the pandemic.

## Conclusions

Our results show two relatively simple ways of comparing the reported evolution of COVID cases across states, regions, or countries with heterogeneous testing. We used test-elasticities or a growth decomposition between test and PTR growth. Data required for our estimates are readily available to public health officials through COVID-19 data repositories; data and code for our analyses are also available. As the pandemic unfolds and more consistent data hopefully become available [10], we hope these results can help support evidence-based decisions by public health officials.

## Data Availability

All data analyzed in this study are publicly available at the following sites:
1. Centers for Medicare and Medicaid Services. Health expenditures by state of residence, 1991-2014. Centers for Medicare & Medicaid Services, Baltimore, MD. 2017.
2. European Centre for Disease Prevention and Control. Data on the geographic distribution of COVID-19 cases worldwide. eCDC, Solna, Sweden. 2020.
Institute of Health Metrics and Evaluation. 2020. Global Health Data Exchange. IHME, University of Washington, Seattle, WA.
3. The COVID Tracking Project. State by state data and annotations. USA. 2020.
4. World Health Organization. Global Health Expenditure Database. WHO, Geneva. 2020. Accessed April 2020.

https://go.cms.gov/2KkMk0f

https://bit.ly/2XWhCm2

http://ghdx.healthdata.org/

https://covidtracking.com/data

https://apps.who.int/nha/database

## Declarations

### Funding

This research was partially supported by the Millennium Science Initiative of the Ministry of Economy, Development and Tourism, Government of Chile, grant “Millennium Nucleus for Collaborative Research in Antimicrobial Resistance”; by FONDECYT Grant 11191206, and research support provided by CEDEUS, ANID/FONDAP 15110020.

### Conflicts of interest

The authors declare no conflict of interest.

### Availability of data and material

All data analyzed in this study are publicly available at the following sites:

Centers for Medicare and Medicaid Services. Health expenditures by state of residence, 1991-2014. Centers for Medicare & Medicaid Services, Baltimore, MD. 2017. https://go.cms.gov/2KkMk0f. Accessed April 2020.

European Centre for Disease Prevention and Control. Data on the geographic distribution of COVID-19 cases worldwide. eCDC, Solna, Sweden. 2020. https://bit.ly/2XWhCm2.

Institute of Health Metrics and Evaluation. 2020. Global Health Data Exchange. IHME, University of Washington, Seattle, WA. http://ghdx.healthdata.org/.

The COVID Tracking Project. State by state data and annotations. USA. 2020. https://covidtracking.com/data. Accessed April 2020.

World Health Organization. Global Health Expenditure Database. WHO, Geneva. 2020. https://apps.who.int/nha/database. Accessed April 2020.

### Code availability

Analysis was performed using Stata; code are available from the authors upon request.

## Acknowledgments

This research was partially supported by the Millennium Science Initiative of the Ministry of Economy, Development and Tourism, Government of Chile, grant “Millennium Nucleus for Collaborative Research in Antimicrobial Resistance”; by FONDECYT Grant 11191206, and research support provided by CEDEUS, ANID/FONDAP 15110020. We are grateful to Laura Marshall for excellent research assistance.

## Notes

### Competing Interest Statement

The authors have declared no competing interest.

